# Principles and Practice of SARS-CoV-2 Decontamination of N95 Masks with UV-C

**DOI:** 10.1101/2020.06.08.20125062

**Authors:** Thomas Huber, Olivia Goldman, Alexander E. Epstein, Gianna Stella, Thomas P. Sakmar

## Abstract

A mainstay of personal protective equipment (PPE) during the COVID-19 pandemic is the N95 filtering facepiece respirator. N95 respirators are commonly used to protect healthcare workers from respiratory pathogens, including the novel coronavirus SARS-CoV-2, and are increasingly employed by other frontline workers and the general public. Under routine circumstances, these masks are disposable, single-use items, but extended use and reuse practices have been broadly enacted to alleviate critical supply shortages during the COVID-19 pandemic. While extended-time single use presents a low risk of pathogen transfer, repeated donning and doffing of potentially contaminated masks presents increased risk of pathogen transfer. Therefore, efficient and safe decontamination methods for N95 masks are needed to reduce the risk of reuse and mitigate local supply shortages. Here we review the available literature concerning use of germicidal ultraviolet-C (UV-C) light to decontaminate N95 masks. We propose a practical method for repeated point-of-use decontamination using commercially-available UV-C crosslinker boxes from molecular biology laboratories to expose each side of the mask to 800–1200 mJ/cm^2^ of UV-C. We measure the dose that penetrated to the interior of the respirators and model the potential germicidal action on SARS-CoV-2. Our experimental results, in combination with modeled data, suggest that a two-minute UV-C treatment cycle should induce a >3-log-order reduction in viral bioburden on the surface of the respirators, and a 2-log order reduction throughout the interior. The resulting exposure is 100-fold less than the dose expected to damage the masks, facilitating repeated decontamination. As such, UV-C germicidal irradiation (UVGI) is a practical strategy for small-scale point-of-use decontamination of N95s.

## Introduction

The recent SARS-CoV-2 outbreak has created a worldwide shortage of personal protective equipment (PPE) which leaves healthcare workers dangerously ill-equipped to aid COVID-19 patients (Ranney et al., 2020). Respiratory viruses such as SARS-CoV-2 can potentially be transmitted through contact, respiratory droplets, and, under certain circumstances, aerosols. WHO recommends droplet and contact precautions for people caring for COVID-19 patients and recommends airborne precautions, such as N95 masks, for circumstances where SARS-CoV-2 aerosol particles are generated (WHO, 2020)

N95 masks are filtering facepiece respirators (FFRs) that have at least 95% efficiency for filtering airborne particles (size around 300 nm) and meet the air filtration rating of the U.S. National Institute for Occupational Safety and Health (NIOSH) classification for filtering respirators. Equivalent standards of N95 are FFP2 (European Union), KN95 (China), DS/DL2 (Japan), and KF94 (South Korea) (Liao et al., 2020). N95 filtering facepiece respirators for use by the general public in public health medical emergencies are intended for over-the-counter (OTC) use (FDA, 2007). A model for a hypothetical influenza pandemic in the US suggested that several billion N95 masks would be required – a number that would lead to severe supply shortages (Carias et al., 2015). Current guidelines during the COVID-19 pandemic suggest limiting the use of N95 masks to healthcare personnel, while providing low-tech face masks for the general public (FDA, 2020a).

Typically, N95 masks are discarded after each use to prevent exposure of infectious material on the respirator to others or the wearer, and extended single use is recommended rather than re-use in the case of pathogens where contact transmission is possible (CDC, 2020a, b). However, due to shortages of PPE, many healthcare workers are not only wearing N95 masks for extended periods of time, but also reusing them (Kobayashi et al., 2020).

There are challenges in maintaining the physical and electrostatic properties of N95 masks after certain decontamination methods. The multi-layer sandwich structure of N95 masks, like the 3M model 8210 (Figure 1), consists of an outer layer (Coverweb) which faces the environment, middle layers (Filters 1 and 2), and an inner layer (Shell) which faces the user (Fisher et al., 2010). Filters 1 and 2 are the internal filtering medium (IFM), the key material of the N95 mask. The IFM is a proprietary Brownian filtration media which traps particles that collide with a specially treated surface, which is made of electrostatically charged meltblown polypropylene microfibers. These microfibers have cross-sectional diameters in the range of 2 to 10 μm and cross each other to form a 3D porous structure, with up to 90% porosity (Liao et al., 2020). The microfibers are electrostatically charged to attract and trap particles and increase filtration efficiency without impeding air flow (Barrett and Rousseau, 1998). Decontamination methods with 75% alcohol or a chlorine-based solution dramatically reduce the filtration performance of N95 masks, likely due to breakdown of the electrostatic charge on the filter material, and are not recommended (Liao et al., 2020). On the other hand, ultraviolet germicidal irradiation (UVGI) has been shown to efficiently inactivate pandemic influenza virus applied as aerosols or droplets on several different N95 masks (Heimbuch et al., 2011; Mills et al., 2018).

**Figure 1.**
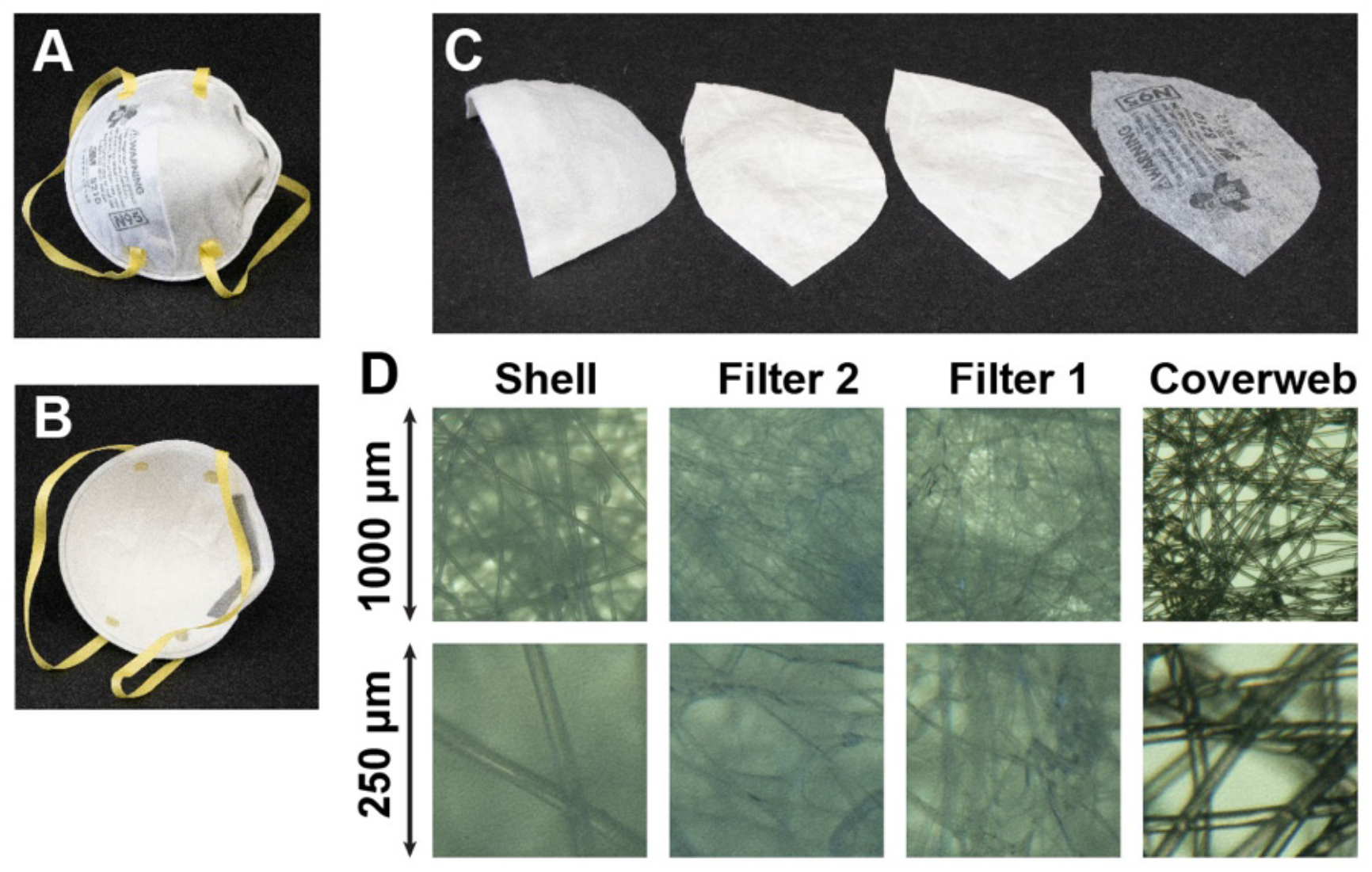
The multi-layer sandwich anatomy of the N95 mask using the 3M Company 8210 mask as an example. (A) Environmental interface. (B) User interface. (C) From *left* to *right;* inner layer (Shell), middle layers (Filter 2 and Filter 1), and outer layer (Coverweb). (D) Light microscope images of the four layers, with lower row at four-fold higher magnification.

Recent work has optimized or implemented UVGI (Lowe, 2020), VHP (Grossman et al.) and moist heat (Anderegg et al., 2020) for large-scale decontamination of N95s in hospitals. However, there is a need for inexpensive, widely available tools for small-scale mask decontamination. We demonstrate that UV germicidal irradiation represents a powerful tool for small-scale decontamination of N95 masks. To justify a minimum irradiation dosage for safe decontamination, we model the germicidal action of UV irradiation in the filter material of the N95 mask. We show that N95 decontamination can be achieved with commercially-available UV crosslinker devices commonly found in molecular biology laboratories. Similar low-cost UV boxes designed for point-of-use N95 mask decontamination could be rapidly produced and distributed worldwide to alleviate local critical shortages of N95 masks, such as during the ongoing COVID-19 pandemic.

## Results

### Physical dimensions

We determined the thickness and areal density for each of the materials of the four layers of the 3M 8210 N95 mask (see Figure 1). The Coverweb, Filter 1, Filter 2 and Shell layers are respectively 0.20 mm, 0.36 mm, 0.41 mm and 0.90 mm thick, and have areal densities of 4.2 mg/cm^2^, 5.1 mg/cm^2^, 5.5 mg/cm^2^ and 16.4 mg/cm^2^. Light microscopy revealed that the fibers within each layer consists are 16 μm (95% CI, 12 to 20) thick in the Coverweb, 5.5 μm (95% CI, 3.9 to 7.1) thick in Filter 1, 2.8 μm (95% CI, 0.7 to 4.8) thick in Filter 2, and 21 μm (95% CI, 19 to 24) thick in the Shell. The packing volume fraction of the microfibers can be approximated as *σ/ρd*_0_, where *σ* is the areal density, *ρ* is the density of the bulk polymer, and *d*_0_ is the layer thickness. With a density for neat polyester as 1.68 g/cm^3^ (for polyethylene terephthalate) and for neat polypropylene as 0.9 g/cm^3^, we estimate the packing volumes fractions as 11%, 15%, 16%, and 13% for Shell, Filter 2, Filter 1, and Coverweb, respectively. The relative internal surface area of the microfibers compared to the macroscopic area of the filter layer can be approximated as 4*σ/ρd* where *σ* is the areal density, *ρ* is the density, and *d* is the fiber thickness. We estimate the relative internal surface areas as 19 for the Shell, 87 for the Filter 2, 41 for the Filter 1, and 6 for the Coverweb.

### Penetration of UV-C light into N95 mask layers

A central question is how much germicidal UV light penetrates the different layers of an N95 mask. The optical transmittance of the mask layers is hard to quantify for short-wavelength UV-C light, especially for the optically dense Shell layer of the N95 mask studied here. We used three complementary methods, spectrophotometry, radiometry, and densitometry, to determine that the optical transmittance of the Shell layer is only 0.1% of the incident irradiance.

Figure 2 shows the spectrophotometric optical transmittance of the different layers of the 3M 8210 N95 mask. The transmittance of the two middle layers (Filter 1 and Filter 2) in the UV-C range is higher than that of the outer layer (Coverweb) and the inner layer (Shell). The Shell and Coverweb layers transmit little UV-C light; they are made of polyester, which is an aromatic polymer and therefore absorbs UV. By contrast, Filters 1 and 2 are made of polypropylene, which has low specific UV absorption besides losses due to light scattering.

**Figure 2.**
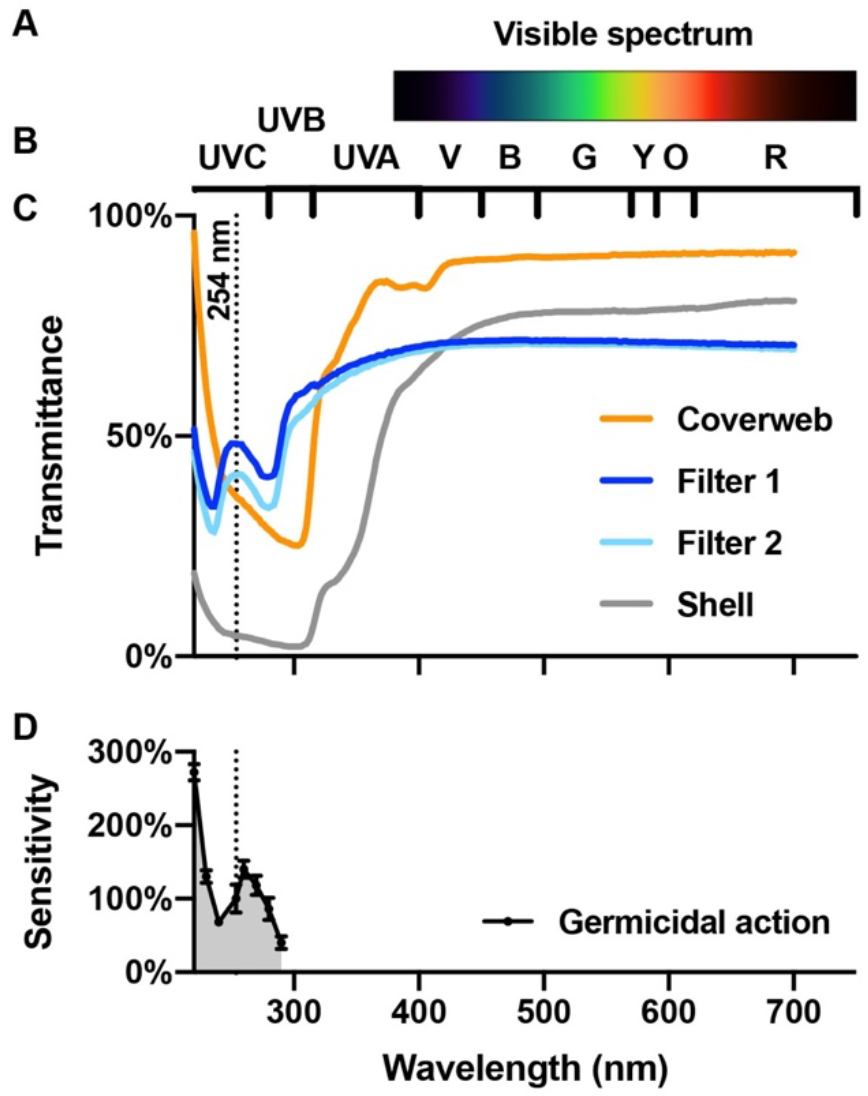
Optical transmittance of N95 mask materials and spectral sensitivity of viral RNA. (A) Colors of the visible spectrum. (B) Wavelength ranges corresponding to different colors (R, red; O, orange; Y, yellow; G, green; B, blue; V, violet) and ultraviolet A, B and C (UV-A, UV-B, UV-C). (C) Optical transmittance of the four layers of the 3M 8210 N95 mask (Coverweb, Filter 1, Filter 2 and Shell). The dotted line indicates the location of the strong 254-nm line of the spectrum of a low-pressure mercury vapor lamp. (D) Germicidal action spectrum shows the wavelength-specific sensitivity of single-stranded RNA viruses. The sensitivity is calculated as the inactivation rate constants for viral infectivity normalized to the value at 254-nm, as determined for the MS2 bacteriophage (Beck et al., 2015).

Since the stray light performance of the spectrophotometer limits the accuracy of the measurements by systematically overestimating transmittance, we employed UV-C radiometry as an alternative approach to measure the spectral transmission of N95 mask materials. The transmittances are 15%, 27%, 21% and 0.11% for the Coverweb, Filter 1, Filter 2 and Shell, respectively.

### Germicidal UV sources

The 254-nm line of the mercury-vapor emission spectrum is the predominant line of a low-pressure mercury-vapor lamp manufactured with a soda-lime glass bulb, which absorbs the 184-nm line. The 254-nm UV-C light efficiently inactivates single-stranded RNA viruses as seen in the germicidal action spectrum (Fig. 2D) (Beck et al., 2015). Recently, germicidal UV light emitting diodes (LEDs) became commercially available that emit in the 260 to 280 nm range.

Figure 3 shows a semi-logarithmic plot of the dose-dependent inactivation of SARS-CoV (Kariwa et al., 2004, 2006). The endpoint dilution assay quantifies the virus titer by serially diluting the treated virus stock, inoculating Vero E6 cells with the serial dilutions, observing the cytopathic effect under a microscope after 48 hours incubation to quantify the percentage of cell death, and calculating the fifty-percent tissue culture infective dose (TCID_50_). We model the data as a double-exponential decay function that enables interpolation of the data points in a meaningful manner. The fast component is 98.6%±0.4% and has a UV-C sensitivity of 0.522±0.040 cm^2^/mJ. The slow component is 1.4%±0.4% and has a sensitivity of 0.066±0.003 cm^2^/mJ. We interpret the slow component as virus trapped at the bottom of the sample and the fast component as freely diffusing virus particles that sample the available volume. Therefore, the fast rate constant is averaged depth dependent UV-C dosage scaled by the sensitivity of the virus. The slow component is the UV-C dosage reaching the bottom. The final plateau with a value of 17±2 TCID_50_/mL value, which corresponds to a fraction of 5×10^-7^, corresponds to the detection limit of the assay, which is likely due to cytopathic effects of protein components of the virions that are not inactivated by the UV-C irradiation, since the assay in principle does not discriminate cytopathic effects from true infectivity as other assays.

**Figure 3.**
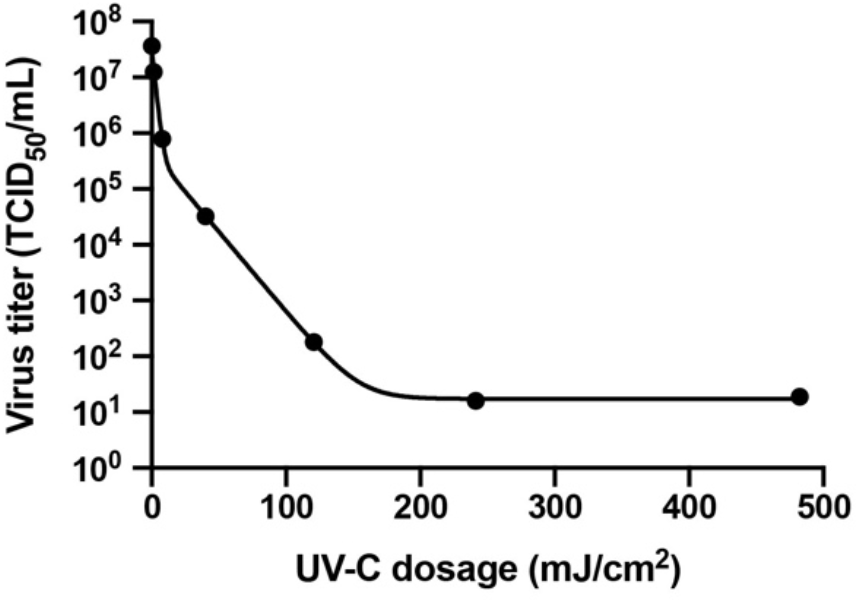
UV-C dose-dependent inactivation of SARS-CoV. We fitted the original experimental data (Kariwa et al., 2004, 2006) to a double exponential model.

### UV-C light sensitivity of SARS-CoV

The sensitivity of RNA viruses depends on several factors, such as genome size and base composition (Kowalski et al., 2009). Moreover, the UV-C dose-dependent inactivation of viruses is known to deviate from a simple first-order decay model (McDevitt et al., 2007). In absence of detailed data on the novel coronavirus SARS-CoV- 2, we use literature data for the closely related virus SARS-CoV (Kariwa et al., 2004), as the UV-C sensitivity of related viruses is roughly constant after accounting for genome size (Lytle and Sagripanti, 2005).

### Modeling the UV-C light distribution in the N95 mask

The relationship of the total optical transmittance and thickness for a scattering and absorbing media should follow an exponential relationship, in which an attenuation coefficient replaces the absorption coefficient of the Beer-Lambert law for non-scattering media. It is based on integrating the time-dependent photon diffusion equation often used to model light penetration into scattering media (Chen et al., 2019). To calculate the attenuation coefficient from the total transmission, we need the actual thickness of each layer.

We use the radiometric UV-C transmittance data to model the depth-dependent UV-C dosage in an N95 mask. The absorbances of the four layers calculated from the transmittance data are 2.68, 0.85, 0.68 and 0.66 for the Shell, Filter 2, Filter 1 and Coverweb, respectively. We model the depth-dependent cumulative absorbance of all layers as a piece-wise linear function corresponding to the sum of the absorbances, with one function from each side. These depth-dependent absorbance functions allow calculation of the depth-dependent transmittance, and therefore the local UV-C dosage.

Figure 4 shows a semi-logarithmic plot with three curves corresponding to the local UV-C dosage as a function of the relative position in the four layers of the N95 mask (Shell, Filter 2, Filter 1, and Coverweb) for three cases of illumination. The first case is illumination from the inside surface of the mask. The orange curve shows the local dosage for illumination from the user-facing inside of the mask with a surface UV-C dosage of 1000 mJ/cm^2^. Due to the high absorbance of the polyester material of the Shell layer, the local dosage rapidly drops to 2 mJ/cm^2^ at the interface between the layers Shell and Filter 2. It further drops to 0.29 mJ/cm^2^ at the interface between Filter 2 and Filter 1, and 0.06 mJ/cm^2^ at the interface between Filter 1 and Coverweb. The residual local dosage at the exit on the other side of the mask is only 0.014 mJ/cm^2^. The second case is illumination from the outside surface of the mask. The green curve shows the local dosage for illumination from the environment-facing outside of the mask with a surface UV-C dosage of 1000 mJ/cm^2^. The dosage drops to 220 mJ/cm^2^ at the interface between Coverweb and Filter 1, to 46 mJ/cm^2^ at the interface between Filter 1 and Filter 2, and to 6.5 mJ/cm^2^ at the interface between Filter 2 and Shell. The exit dosage at the inside surface is 0.014 mJ/cm^2^. The third case is for illumination from both sides of the mask. The black curve shows the local dosage for illumination from both sides of the mask with a surface UV-C dosage of 1000 mJ/cm^2^, each. The local dosage is 8.6 mJ/cm^2^ at the Shell to Filter 2 interface, 46 mJ/cm^2^ at the Filter 2 to Filter 1 interface, and 220 mJ/cm^2^ at the Filter 1 to Coverweb interface. The minimum of the black curve is in the core of the Shell material, close to the Filter 2 layer, where the local dosage is 7.4 mJ/cm^2^. Therefore, the core of the N95 mask receives 270-fold less UV-C light as compared to total surface exposure of 2000 mJ/cm^2^. Notably, a previous attempt to estimate the reduction of UV-C light in the interior of the 3M 8210 mask concluded that there was only a 16.4-fold reduction of UV-C dosage (Fisher and Shaffer, 2011), which we conclude is based on an incorrect mathematical treatment of the radiometric observables.

**Figure 4.**
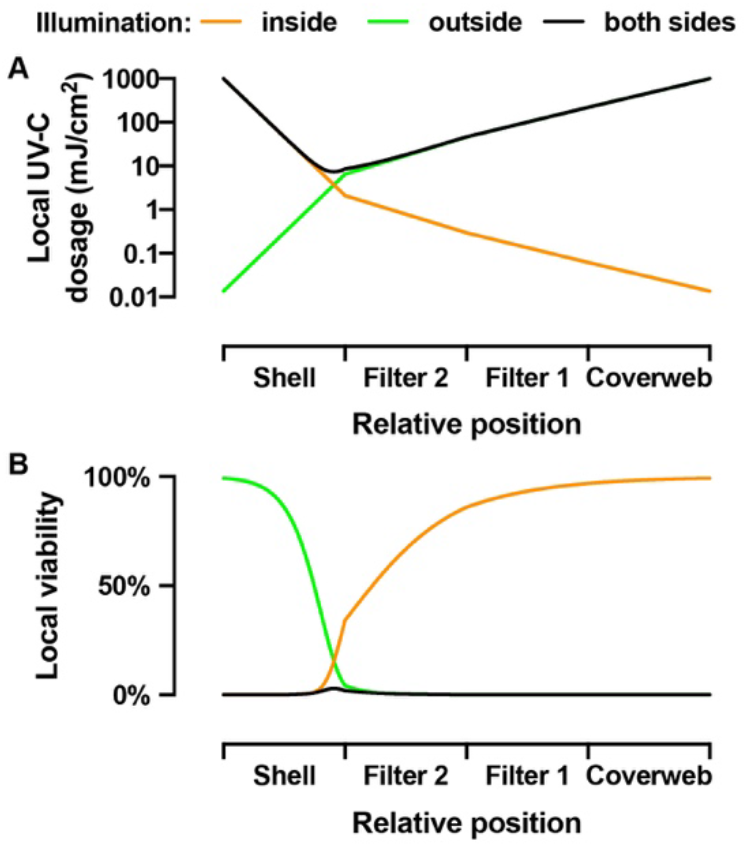
Modeling of the UV-C dosage and virus viability in different layers of the N95 mask. (A) Local UV-C dosage as a function of the relative position inside the four layers of the N95 mask. The three different curves show the local UV dosage for three cases of illumination; illumination from the inside only (orange), illumination from the outside only (green), and illumination from both sides (black) with 1000 mJ/cm^2^ surface UV-C dosage. (B) Local virus viability predicted from the local UV-C dosage distributions for the three cases.

### Modeling the virus viability in the N95 mask

The local UV-C dosage calculated for the three cases of illumination together with the UV-C dose-dependent inactivation of SARS-CoV enables an estimate of the local residual virus viability in an irradiated N95 mask (Figure 4). The orange curve is for the first case with illumination from the inside surface of the mask. The first half of the Shell layer is efficiently decontaminated with 0.07% local viability in the center of the Shell. However, the sharp rise of the local viability to 34% at the interface of Shell and Filter 2 shows that the illumination from the inside of the mask is insufficient to decontaminate the highly porous material of the Shell layer. Note that the optically dense Shell layer has a thickness of 0.90 mm, which is almost as much as the combined thickness of all other layers (Filter 2 is 0.41 mm, Filter 1 is 0.36 mm and Coverweb is 0.20 mm thick). In contrast, the green curve, for the second case where the mask is illuminated from the outside, shows efficient decontamination of the three layers, Coverweb, Filter 1 and Filter 2, with 4.3% local viability at the interface of Filter 2 and Shell, but little or no decontamination of the Shell. For the third case with illumination from both sides, the local viability throughout the mask is very low. The smallest UV-C dosage inside the core of the Shell material results in 3% local virus viability. The integrated local viability yields the averaged virus viability in the mask, which is 0.3% for illumination from each side together of the mask with 1000 mJ/cm^2^ surface dosage.

### Modeling the UV-C dose-dependent virus inactivation

Here we analyze the dose-dependent decontamination levels and the effect of different models of UV-C sensitivity on the achievable degree of decontamination. Previous literature disagrees on different UV-C sensitivity of coronaviruses and other related viruses. Therefore, out of an abundance of caution, we determined the proper UV-C dosage to sufficiently decontaminate N95 masks based on three different models of virus viability as a function of UV-C dose. We also estimated local viability as a function of mask position assuming a 1000 mJ/cm^2^ exposure (Figure 5).

**Figure 5.**
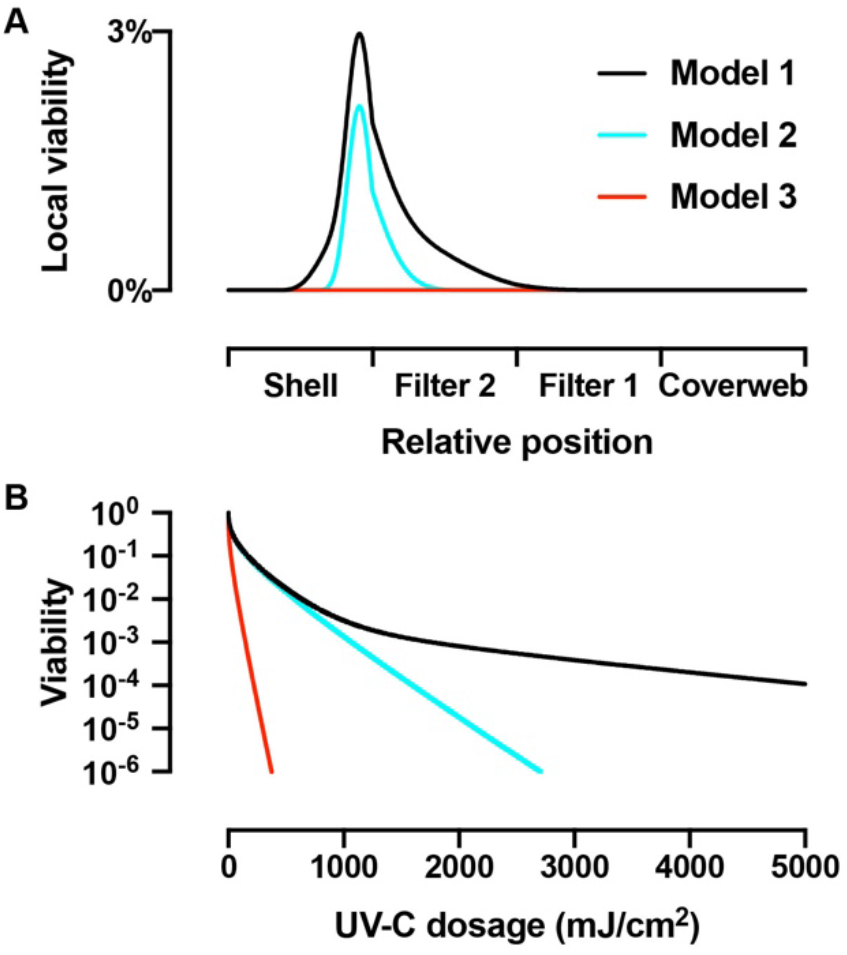
Modeling of the virus viability for three different models of UV-C sensitivity. (A) Local UV-C dosage as a function of the relative position inside the four layers of the N95 mask illuminated from each side together with 1000 mJ/cm^2^ surface dosage. Model 1 (black) uses the empirical double-exponential model to describe the UV-C dose-dependent inactivation shown in Figure 3. Model 2 (cyan) uses an UV-C sensitivity of 0.522 cm^2^/mJ, corresponding to the fast, initial virus inactivation process from Figure 3. Model 3 (red) uses an UV-C sensitivity of 3.77 cm^2^/mJ, which describes the UV-C inactivation of MHV coronavirus aerosols. (B) Total virus viability as a function of total surface dosage equally divided across both sides for the three models.

Based on the data from Figure 3, Model 1 is a data-driven prediction of virus inactivation that accounts for the possibility of virus populations with different UV-C sensitivities (Kariwa et al., 2004, 2006). The predicted local viability (black curves in Figure 4B and Figure 5A) shows the difficulty to inactivate the virus in the core of the Shell and, to a smaller extent, in Filter 2, but the model demonstrates sufficient decontamination to prevent surface transmission for re-use of an N95 mask, despite the 1.4% of the virus population that shows reduced UV-C sensitivity. However, we suspect that the UV-C sensitivity predicted by Kariwa *et al*. may be underestimated due to confounding inner filter effects in the experimental assay (see *“Potential confounds in measurement of viral UV-C sensitivity”* in the discussion).

Therefore, we created Model 2 (cyan curves in Figure 5), which assumes that the major population, consisting of 98.6% of virus, has a UV-C sensitivity of 0.522 cm^2^/mJ based on observations from Darnell et al. (2004). The local virus viability in the Shell is similar in both Models 1 and 2. However, because of the lack of the UV-C resistant minor population in Model 2, more virus is inactivated in the Filter 2 since the UV-C resistant minor population is the main contributor at low percentage values of local viability. Model 3 (red curves in Figure 5) is similar to Model 2, but assumes a higher UV-C sensitivity of 3.77 cm^2^/mJ based the measured UV-C sensitivity murine hepatitis virus (MHV) coronavirus in aerosol (Walker and Ko, 2007). Model 3 suggests complete inactivation of virus in all layers of the mask, including the highly shielded core of the Shell.

We calculated the total virus viability averaged over all layers, as obtained from the integrated local viabilities, for the three different models as a function of surface UV-C dosage (Figure 5B). The curves for Model 1 (black) and Model 2 (cyan) are similar for UV-C dosages up to 1000 mJ/cm^2^, before they diverge. The surface dosages from each side necessary to achieve 2-log order reduction to 1% total viability are 637 mJ/cm^2^ for Model 1 and 575 mJ/cm^2^ for Model 2. For 3-log order reduction to 0.1% total viability, the dosage for Model 1 with 1765 mJ/cm^2^ is already noticeably larger than the 1068 mJ/cm^2^ for Model 2. Model 1 fails to achieve 4-log order reduction for dosages up to 5000 mJ/cm^2^, where the total virus viability reduced to 0.011%. Model 2, on the other hand, reaches 4-log order reduction to 0.01% at 1597 mJ/cm^2^, 5-log order reduction at 2145 mJ/cm^2^, and 6-log order reduction at 2705 mJ/cm^2^. In comparison, the high UV-C sensitivity in Model 3 predicts 3-log order reduction at 148 mJ/cm^2^ and 6-log order reduction at 375 mJ/cm^2^. Model 3 shows robust UV-C sensitivity and corresponds to a 3-log reduction with notably lower UV dosage compared to Model 1 and Model 2.

## Discussion

In this section, we review literature and outline principles and points of consideration when implementing UVGI decontamination of N95 masks. The optimization of the UV-C dosage for efficient decontamination of a mask depends on several design parameters. What degree of reduction of virus viability is desired? Is it necessary to decontaminate interior layers of the mask that are inaccessible for direct contact? Does the applied UV-C dosage negatively affect the performance of the mask? Is it practical to deliver the necessary dosage?

### N95 mask filtration efficiency for aerosolized viruses

Aerosolized MS2 virions permeate N95 masks with a peak permeability exceeding 5% for particles with approximately 50 nm diameter (Balazy et al., 2006; Eninger et al., 2008), but the permeability is below 1% for aerosolized MS2 at 500 nm average particle diameter (Gardner et al., 2013). Studies have demonstrated that N95 masks trap MS2 virions in small aerosol particles (with 141 nm median diameter) preferentially in the middle layer, whereas virions in larger aerosol particles (median diameter 492-nm) are preferentially trapped in the outer layer (Fisher et al., 2009). The only study that used aerosolized adenovirus and influenza virus found levels of permeability comparable with the results from the MS2 model virus, even though the MS2 virion (27.5 nm diameter) is smaller than influenza virus (80 to 120 nm diameter) or SARS-CoV-2 (50 to 200 nm diameter) (Zuo et al., 2013). This suggests that the size of the aerosol particles carrying virions, not the size of the virions themselves, are the important factor for the efficacy of N95 mask’s mechanism of action.

Hospital samples of SARS-CoV-2 aerosols collected during the outbreak in Wuhan, China showed viral RNA present in a distribution of different sized aerosols with multiple peaks: one in submicron region (particle diameter of 250 nm to 1 μm) and another in the supermicron region (particle diameter larger than 2.5 μm), but also in even smaller particles (range 10 nm to 250 nm) (Liu et al., 2020). It is noteworthy that live virions in environmental aerosols are key to transmission, and it is difficult to quantify compared to viral RNA alone. Quantification from Liu et al. is based on RT-qPCR assays of viral RNA instead of culturing viable virions (Tellier et al., 2019). Based on studies using a similar distribution of different-sized MS2 aerosol particles, a large fraction of the supermicron and submicron SARS-CoV-2 particles would be trapped in the outer layer of N95 masks, whereas the smaller particles should get trapped in the microfiber material of the middle layer.

### Routes of transmission via N95 mask extended use/reuse

For decontamination of N95 masks, it is important to consider which parts of the multilayer structure provide potential viral transmission risk. Risk of contact transmission is due to handling the contaminated surface of a mask (CDC, 2020b). It is also possible that virus could be re-aerosolized from the mask due to the airflow from breathing, coughing or sneezing. A study of simulated coughing through an N95 mask contaminated with the MS2 virus demonstrated that only a small percentage of virus was released (Fisher et al., 2012). However, SARS-CoV-2 RNA was present at high levels in large particle aerosols in hospitals during the COVID-19 outbreak, which raises concerns that larger particles are more efficiently re-aerosolized under similar conditions (Qian et al., 1997). (Liu et al., 2020). Our results demonstrate that inner layers of the N95 mask will inevitably receive different amounts of dosage than outer layers. Decontamination methods must consider efficacy of inactivating virions most importantly on the masks’ surface, as well as on the inner layers of the mask.

### Potential confounds in measurement of viral UV-C sensitivity

The prediction and measurement of the sensitivity of novel viruses to germicidal UV light is an important and difficult problem (Kowalski et al., 2019; Kowalski et al., 2009; Lytle and Sagripanti, 2005). Several studies have investigated UV-C sensitivity for coronaviruses, including SARS-CoV (Darnell et al., 2004; Duan et al., 2003; Kariwa et al., 2004, 2006; Liu et al., 2003) and SARS-CoV-2 (Fischer et al., 2020).

A confound in these studies is that the virus preparations are made in DMEM including 10% FBS and other additives, such as antibiotics and antifungals. The high protein content of FBS results in a high absorbance at 254 nm. We determined that 10% FBS contributes 1.54±0.04 and DMEM contributes 1.12±0.02 absorbance at 254 nm. The absorbance is about 2.66±0.04 per 1-cm pathlength with about 58% contribution from FBS and the rest from DMEM. Therefore, even shallow layers of virus suspension will have a high absorbance that results in substantial inner filter effects that reduce the available UV-C dosage in a depth-dependent fashion. (For clarity, we would like to note that “inner filter effect” is a principle in fluorescence spectroscopy and that has no relation in concept to N95 mask filters.)

These inner filter effects likely explain the discrepancies between two studies with SARS-CoV, where the reduction of the filling height of the virus suspension from 1 to 0.25 cm resulted in 40-fold increase of UV-C sensitivity from 0.012 to 0.477 cm^2^/mJ (Darnell et al., 2004; Kariwa et al., 2004). The UV-C sensitivity of a different coronavirus, murine hepatitis virus (MHV), in aerosol form was 3.77 cm^2^/mJ (Walker and Ko, 2007), which is even 8-fold higher than SARS-CoV in the shallow solution. Since the small diameter of the aerosol droplets will have negligible inner filter effects, UV-C sensitivities measured in aerosol is most applicable, although to perform these experiments on additional viral species is prohibitive due to requisite biosafety measures.

UV-C inactivation of the Berne virus or equine torovirus, closely related to coronaviruses, has been measured in very shallow preparations of about 0.05-cm path length (Weiss and Horzinek, 1986). The corresponding UV-C sensitivity is 3.24 cm^2^/mJ, which is very close to that for the MHV aerosols, additionally supporting that the high UV-C sensitivity inferred from the aerosols and is most closely reflected by values determined by solutions with minimal inner filter artifacts. Future studies of the UV-C sensitivity of SARS-CoV-2 should carefully address the issue of UV attenuation due to the sample media composition, for example, by using very thin solution layers or by using virus purified in low UV-absorbing buffers.

### Potential methods for validating N95 decontamination

In addition to developing models that predict UV-C penetration into masks and inactivation of SARS-CoV-2, the reduction in viral bioburden in N95 masks could potentially be measured directly. Oral et al. demonstrated the effectiveness of VHP decontamination by applying SARS-CoV-2 to the surface of an N95 mask and assessing the effect of treatment through quantifying extracted of viable virus from mask sections (Oral et al., 2020). Future work could employ this technique to assess the ability of UV-C to affect viability of SARS-CoV-2. However, this method of measuring bioburden reduction has a key limitation; the static application of the virus to the respirator surface does not accurately model the penetration of aerosolized virus deeper into the respirator. Aerosolization of the virus prior to exposure is prohibitively dangerous, requiring a biosafety level 4 (BSL-4) laboratory.

### Small-scale implementation of UVGI for N95 masks

Many groups have focused on large-scale chambers to accommodate up to 27 masks in one cycle with two planar 12 UV-C bulb arrays (Hamzavi et al., 2020; Schnell et al., 2020). However, biosafety cabinets and even medium-scale UVGI boxes that hold 6 masks demonstrate variability in UV dosage as a result of distance, angle of incidence from light source, and orientation and shape of the masks (Baluja et al., 2020; Card et al., 2020). The higher UV-C doses needed to compensate for this variability and ensure decontamination may increase the risk of compromising masks’ structural integrity. By contrast, a single-mask decontamination system can ensure that a controlled, carefully measured dose UV-C dose is delivered to the mask. In addition, such a system would be cheap and practical for point-of-care decontamination outside of hospital settings.

We used a commercial UVP Crosslinker that has 5x 8W 254-nm mercury vapor bulbs in an enclosure. A microprocessor controls the UV-C exposure and stops the irradiation when the desired energy is delivered. Since the bulbs are only at the top of the enclosure, we needed to use two sequential exposures, flipping the mask upside-down in-between them. The need to flip the masks adds another handling step with the risk of viral contamination. Point-of-use UV-C decontamination of N95 masks could prove useful to frontline workers, such as firefighters, transit workers, and grocery store employees, who would need to decontaminate small numbers of respirators and would lack access to the bulk respirator decontamination systems now being implemented in hospitals.

### Photodegradation of the masks

The photodegradation of polypropylene filaments by UV-C irradiation has been studied with mechanical strength tests and Fourier transform infrared (FTIR) spectroscopy, which has demonstrated the formation of alcohols, peroxides, ketones, aldehydes, carboxylic acids, and anhydride reaction products (Aslanzadeh and Kish, 2010; Mahmoodabadi et al., 2018). The initiation of photo-oxidation reaction cascades of polypropylene, especially at wavelengths longer than 290 nm, is thought to originate from chromophoric groups, such as hydroperoxides, which in turn are formed in an autocatalytic process (Feldman, 2002). The photodegradation of the polyester poly(ethylene terephthalate) involves photo-oxidative reactions leading to chain scission and the generation of carboxyl end groups (Fechine et al., 2004; Hurley and Leggett, 2009).

Lindsley et al. studied the effect of germicidal UV irradiation on the mechanical stability and filter performance of N95 mask materials (Lindsley et al., 2015). For four tested N95 models, the mechanical strength of one or more layers was significantly reduced at exposure doses higher than 120000 mJ/cm^2^ sequentially applied to each side. While they observed only minor changes in particle penetration and flow resistance as a result, they did not determine whether changes in the material would affect the ability of a mask to conform to the user’s face. Although the dose required for decontamination is 100-fold lower (1000-2000 mJ/cm^2^), we wanted to confirm that UV-C irradiation would not affect fit of N95 respirators even after multiple decontamination cycles.

We exposed two 3M model 8210 N95 respirators to repeated doses of UV-C (with either 5000 or 10000 mJ /cm^2^ exposure doses per side). Before and after each cycle, the masks were donned by a volunteer, and the quantitative fit factor was measured using a TSI PortaCount Pro Plus. The quantitative fit factor is defined as the ratio of concentrations of 0.02-1 μm particles outside and inside the mask, and is decreased both by particle penetration and poor fit. For N95 masks and other disposable respirators, the PortaCount Pro can measure a maximum fit factor of 200 (which would correspond to the mask filtering at least 99.5% of particles). One mask was subjected to fourteen exposure cycles (5000 mJ/cm^2^ per cycle per side), and the other to one long exposure (20000 mJ /cm^2^ exposure doses per side) followed by six shorter cycles (10000 mJ/cm^2^ per side) giving 80000 mJ/cm^2^ total dosage per side. Both tested masks maintained a fit factor of 200 during both normal breathing and deep breathing throughout for exposures up to 50000 mJ/cm^2^ total dosage per side, which would correspond to fifty decontamination cycles with 1000 mJ/cm^2^ each per side.

These results differ from a separate study using quantitative fit factor measurements of N95 masks decontaminated by UV-C. A significant reduction in quantitative fit factor was observed, although not below sufficiency, after a dose of approximately 18000 mJ/cm^2^ via a biosafety cabinet (Smith et al., 2020). This would correspond to nine two-minute decontamination cycles in the “Local UV” box. We note that this study used different models of N95 masks (1860, Aura 1870+, industrial 8511) manufactured by 3M, although no data were provided investigating differential effects of UV-C for each model. Another study of the effect of UV-C irradiation on the fit test performance of N95 masks did not have data for the 3M 8210 mask due to the current supply shortage (Price et al., 2020). Further testing is needed on other N95 models to determine the number of two-minute decontamination cycles that they can withstand without compromise.

We did observe that the UV-C irradiation of our N95 masks resulted in a slight odor of the masks that appeared to originate from the polyester material of the Shell layer, since the odor was similar upon irradiation from both sides or only the inside, where all the light is absorbed by the Shell layer. The odor is weak and dissipates rapidly, but nevertheless might reduce the acceptance of the UV-C decontamination of N95 masks for sensitive individuals. Others have described nutty/smoky odor of masks after UV-C decontamination and recommended to allow for off-gassing time when feasible (Schnell et al., 2020). Odor has been reported for moist heat decontamination methods as well (Viscusi et al., 2011).

### Present and future importance of N95 emergency reuse

Epidemiological reports indicate that the number of infectious disease outbreaks tripled between 1980 and 2010 (Smith et al., 2014), indicating a high likelihood of future moments of acute need for PPE such as N95 masks. It is difficult to prepare or stockpile N95 masks supplies in anticipation of this, given that current models of N95s can have a shelf life of 5 years, and decrease in effectiveness after their manufacturer-designated expiration date (CDC, 2020a, b). Furthermore, challenges in increasing N95 production may not allow demand to be reached quickly enough during critical early moments of the spread of a disease and suggests that emergency N95 reuse is a necessary option to maintain. We note that two months after initial shortages of N95s due to COVID-19, demand has still not been met for health care workers treating infected individuals (FDA, 2020b). Therefore, we suggest that decontamination methods, such as UVGI of N95 masks, be considered as a feature of preparedness for times of acute need of PPE.

## Conclusion

Several strategies have been used to decontaminate N95 masks during the COVID-19 pandemic, most of which are being developed for large-scale applications. Here we review the efficacy of UV-C decontamination for N95s, considering factors such as UV transmittance to different layers of the mask, viral sensitivity to UV-C, and potential photodegradation of masks. We also describe the use a UV crosslinker box commonly found in molecular biology laboratories as a practical, point-of-use method for small-scale rapid UV-C decontamination of N95 masks. Such devices assure that a consistent dose of UV-C is applied to the masks, enabling reliable decontamination and repeated reuse without substantial mask photodegradation. Mass production of similar custom low-cost devices could be a cost-effective way of empowering frontline workers, and potentially the general public, to decontaminate masks even if they lack access to a large hospital-based decontamination facility.

## Data Availability

All data presented in this manuscript will be made available upon request from.

## Acknowledgements

We thank the Rockefeller University laboratories of Drs. Paul Bieniasz, Frederick R. Cross, Seth A. Darst, Titia de Lange, Elaine Fuchs, Hironori Funabiki, Howard C. Hang, Mary E. Hatten, Sebastian Klinge, Roderick MacKinnon, Paul Nurse, Michael O’Donnell, Charles M. Rice, Michael P. Rout, Agata Smogorzewska, Hermann Steller, and Leslie B. Vosshall for generously providing access to their UV crosslinkers for this project. We thank Gaitree McNab, Frank X. Schaefer and Amy Wilkerson of the Rockefeller University Department of Laboratory Safety and Environmental Health, and in particular Vichelle Filoteo, Eunice Jung and Sachin Kadam for N95 fit testing. We also thank Bryan Baker, Ann H. Campbell, Dr. Daniel Gareau, Dr. Hiroyuki Takai, and Dr. Iltefat Hamzavi.

## Materials and Methods

### UV-C transmittance measurements

The hemispherical transmittance of a scattering medium is usually measured with an integrating sphere. A simple alternative is to use an optical diffusor, such as a opal glass or filter paper behind the sample (Amesz et al., 1961).

The hemispherical transmittance (*T*) can be calculated from the ratio of the transmitted irradiance (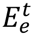) and the incident irradiance (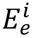):

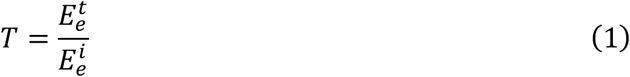

### Transmittance measurement with UV-Vis spectroscopy

To determine the absorbance spectrum of each of the different layers of the N95 mask, we adapted a method normally used for scattering suspensions (Amesz et al., 1961). We placed a piece of Whatman filter paper as a light-diffuser behind the blank and sample cuvette holder in a double-beam UV-Vis spectrophotometer (Perkin Elmer Lambda 800). We taped a piece of the mask material in front of the filter paper in the sample position. In this way, the light- diffuser collected a representative sample of the scattered light penetrating the mask material.

### Transmittance measurement with UV-C radiometry

We used a high-intensity 254-nm UV light source (Analytik Jena US, UVP CL-1000) and a UV-C light meter (General Tools UV254SD) to measure the irradiance with and without a piece of each different layer of the N95 mask in front of the sensor. We calculate the transmittances using equation 1.

We also used a 254-nm UVP Compact UV lamp that facilitates measuring the transmittance of the materials without the need to turn off the lamp when changing the samples. The transmittances were 22% (16 to 29), 21% (16 to 26), 14% (9 to 20) and 0% (0 to 5) for the Coverweb, Filter 1, Filter 2 and Shell, respectively. Values in parentheses are 95% confidence intervals (95% C.I.) from nine repeated sets of measurements, including the dominant contribution from the specified measurement accuracy of the UV254SD meter, which is ±0.082 mW/cm^2^. The lower power of the UVP Compact UV lamp as compared with the UVP CL-1000 source did not allow a non-zero measurement for the optically dense material of the Shell layer. Therefore, we used a densitometric method to quantify more accurately the transmittance of the Shell layer.

### Densitometric UV radiometry

We used photochromic intensity indicators (New UV Intensity Labels, UV Process Supply, Chicago) for direct UV densitometry measurements (Figure 6). These yellow-colored labels change gradually to green when exposed to UV. We determined the UV dose-dependent change in color by exposing a set of labels to increasing UV dosages in the UVP CL-1000 source. We simultaneously recorded the UV irradiance with the UV254SD meter using a one second sampling interval to obtain the actual exposure dosage by numerical integration. We quantified the color change using a flatbed scanner (Epson) and Fiji/ImageJ for image analysis. The red channel of the RGB image showed the largest change of photochromic effect. Exponential fitting of the dose-dependent data with Prism (Graphpad) showed that a dose of 88 mJ/cm^2^ (95% C.I. 77 to 103) yields a half-maximum photochromic effect.

**Figure 6.**
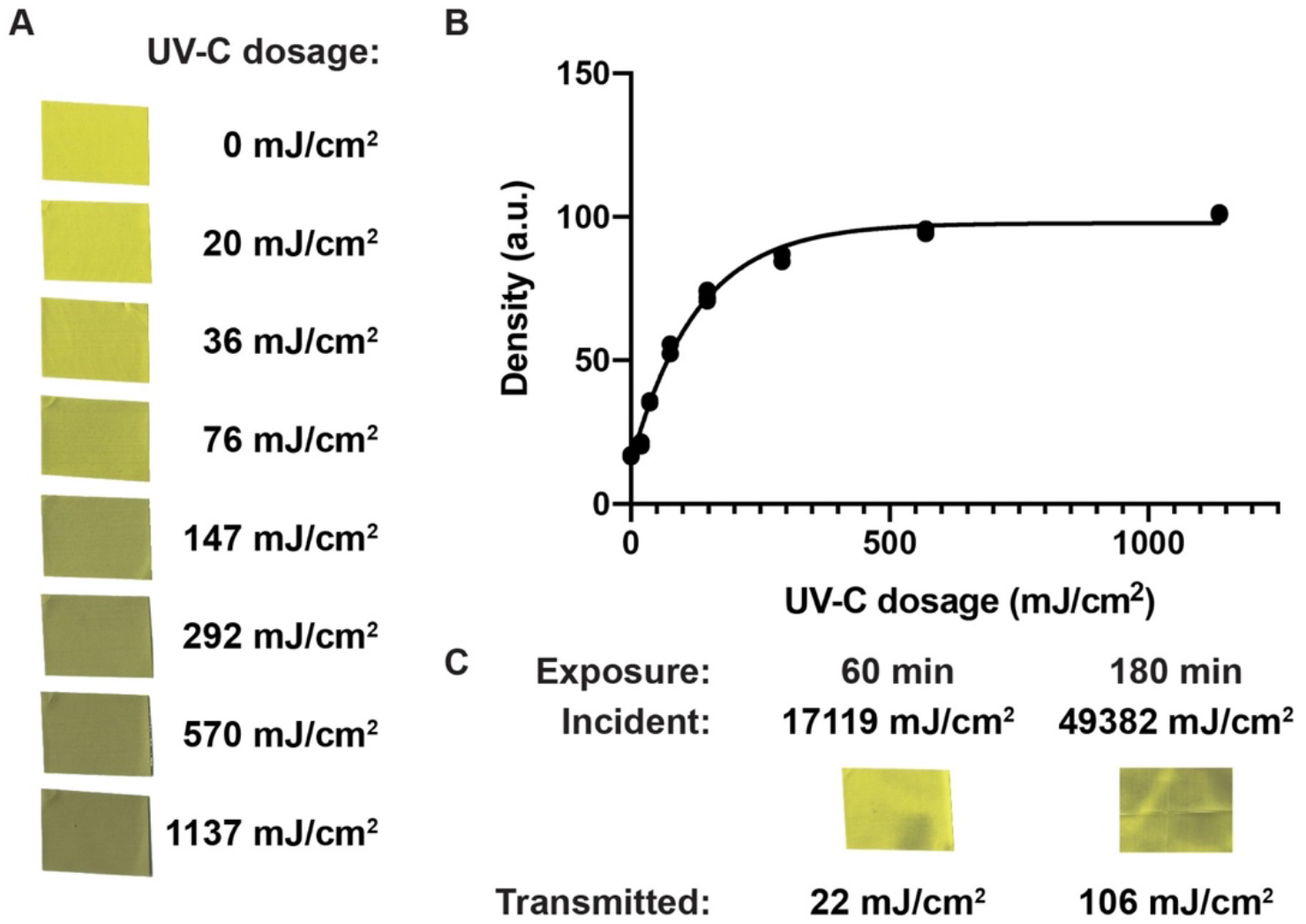
Densitometric UV-C radiometry and transmittance of Shell material. (A) Color change of UV Intensity label stamps upon exposure with increasing UV-C dosage. (B) Exponential fit of the density of the stamps quantified as the mean inverted red channel intensity of the flatbed scanner image. (C) UV intensity labels exposed through the Shell material of the 3M 8210 N95 mask for 60-min and 180-min exposures in the UVP CL-1000 Crosslinker. The incident exposure was quantified by integration of the UVC meter reading sampled in one second intervals. The Transmitted exposure is quantified using the calibration curve from panel (B).

We exposed labels sequentially through the Shell layer material for 60 min in the UVP CL-1000 source. We quantified the color changes of these labels under identical conditions to the labels for the calibration. The 60-min exposure with an integrated UV surface dosage of 17119 mJ/cm^2^ resulted in a color change that corresponds to a transmitted dosage of 22 mJ/cm^2^ (95% C.I. 17 to 26). The ratio of the transmitted and surface dosages gives the total transmission, which is 0.13% (95% C.I. 0.10 to 0.15). We noticed that the color change across the label was uneven, for example with less changes beneath a wrinkle in the Shell layer, due to the manufacturing of the dome-shaped structure.

Next, we exposed an array of intensity labels to map the intensity distribution of the transmitted light. We used a 180-min exposure with 49382 mJ/cm^2^ surface dosage to map the transmitted light intensity distribution in a 5-cm^2^ region under the Shell layer material. The average color change corresponds to a transmitted dosage 106 mJ/cm^2^ (95% C.I. 97 to 116), which gives 0.21% (95% C.I. 0.20 to 0.23). The one standard deviation wide range of color changes corresponds to a range of transmitted dosages from 0.13% to 0.34%. The 95% range of color changes observed for the transmitted light corresponds to a range of transmitted dosages from 0.07% to 5.7%.

### Modeling the UV-C light distribution in the multilayer structure of the N95 mask

The transmittance of the *i*-th layer (*T_i_*) is related to its thickness (*d_i_*) by an attenuation coefficient (*α_i_*):

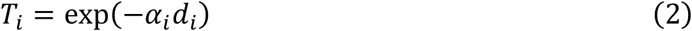

We define a depth-dependent attenuation coefficient as a step function corresponding to the attenuation coefficients of all *n* layers:

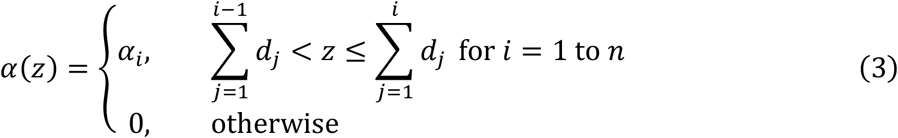

Integration of the depth-dependent attenuation coefficient yields the optical thickness (*τ*) at depth (*d*):

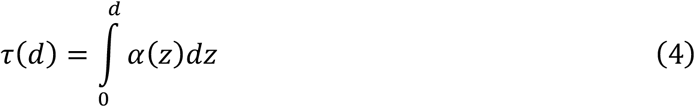

The depth-dependent transmittance (*T*(*d*)) is related to the optical thickness by:

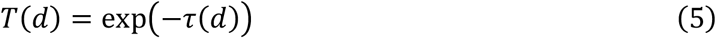

The model neglects the contributions of multiple reflections between the layers of materials, which is small due to the poor reflectance of materials in the short wavelength UV-C region.

We confirmed the validity of our model by control experiments measuring the transmittance of stacks of identical layers of material that give a linear increase of total optical thickness with the number of layers.

The optical thickness in the reverse direction (*τ_R_*) is obtained by integration of the interval starting at the depth (*d*) and ending at the combined thickness of all layers (∑*d_i_*):

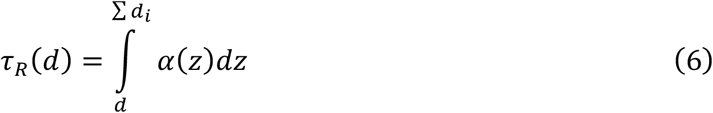

Here we define the depth-dependent transmittance in reverse direction (*T_R_*(*d*)) as:

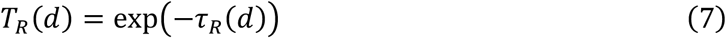

The exposure dosage (*D*) is related to the exposure time (*t*) and the irradiance (*E*_e_)

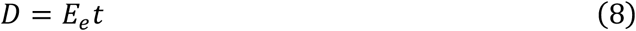

The depth-dependent exposure dosage (*D*(*d*)) is given by the surface exposure in the forward direction (*D*) and the surface exposure in reverse direction (*D_R_*) and the depth-dependent transmittances:

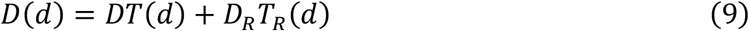

### Modeling the virus viability in the N95 mask

We define the survival (*s*) as ratio of the number of infectious virions after exposure (*n*) and before exposure (*n_0_*):

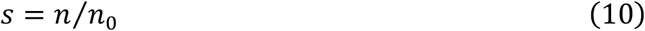

The simplest dose-dependent survival function (*s*(*D*)) for single-hit kinetics of UV inactivation of virus infectivity relates the exposure dosage ((*D*)) to an UV rate constant (*k*):

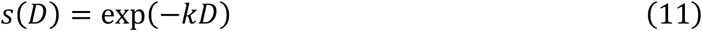

We also use a second model with a secondary population of viruses with a different rate constant (*k′*) that comprises a fraction (*f*) of the total population:

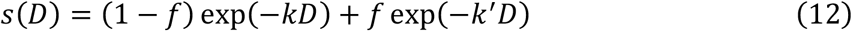

The average survival (〈*s*〉) can be calculated from a depth-dependent density of viruses before exposure (*σ*_0_(*d*)), the survival function and the depth-dependent exposure dosage:

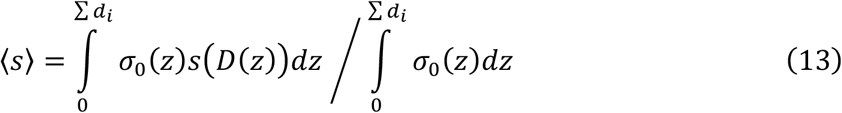

The figures are modeled with a constant value for *σ*_0_(*d*) corresponding to uniform contamination of the mask layers. We also use a unit value for the thickness for each layer (*d_i_* Together these two choices give each layer the same share to the overall virus survival. A more advanced model would include a nonlinear distribution of virus load trapped across the different layers of the mask, based on the adsorption kinetics of the viral particles on the internal surface area of the various layers of different microfiber materials.

